# Plasma proteomics reveals the potential causal impact of extracellular matrix proteins on abdominal aortic aneurysm

**DOI:** 10.1101/2024.09.20.24314065

**Authors:** Samuel Khodursky, Shuai Yuan, Joshua M. Spin, Philip S. Tsao, Michael G. Levin, Scott M. Damrauer

## Abstract

**Background:** Abdominal aortic aneurysm (AAA) is a common and life-threatening vascular disease. Genetic studies have identified numerous associated loci, many potentially encoding plasma proteins. However, the causal effects of plasma proteins on AAA have not been thoroughly studied. We used genetic causal inference approaches to identify plasma proteins that have a potential causal impact on AAA.

**Methods:** Causal inference was performed using two-sample Mendelian randomization (MR). For AAA, we utilized recently published summary statistics from a multi-population genome-wide association (GWAS) meta-analysis including 39,221 individuals with, and 1,086,107 individuals without AAA from 14 cohorts. We used protein quantitative trait loci (pQTLs) identified in two large-scale plasma-proteomics studies (deCODE and UKB-PPP) to generate genetic instruments. We tested 2,783 plasma proteins for possible causal effects on AAA using two-sample MR with inverse variance weighting and common sensitivity analyses to evaluate the MR assumptions. Bayesian colocalization and gene ontology (GO) enrichment analyses provided additional insights.

**Results:** MR identified 90 plasma proteins associated with AAA at FDR<0.05, with 25 supported by colocalization analysis. Among those supported by both MR and colocalization were previously experimentally validated proteins such as PCSK9 (OR 1.3; 95%CI 1.2-1.4; P<1e-10), LTBP4 (OR 3.4; 95%CI 2.6-4.6; P<1e-10) and COL6A3 (OR 0.6; 95%CI 0.5-0.7; P<1e-6). GO analysis revealed enrichment of proteins found in extracellular matrix (ECM, OR 7.8; P<1e-4), some with maximal mRNA levels in aortic tissue. Bi-directional MR suggested plasma level changes were not caused by liability to AAA itself. We then investigated whether variants responsible for expression changes in the aorta also influenced plasma levels and AAA risk. Colocalization analysis showed that an aortic expression quantitative trait locus (eQTL) for COL6A3, and a splicing quantitative trait locus (sQTL) for LTBP4 colocalized with their respective plasma pQTLs and AAA signals (posterior probabilities 0.84 and 0.89, respectively).

**Conclusions:** Our results highlight proteins and pathways with potential causal effects on AAA, providing a foundation for future functional experiments. These findings suggest a possible causal pathway whereby genetic variation affecting ECM proteins expressed in the aortic wall cause their levels to change in blood plasma, influencing development of AAA.

## Introduction

Abdominal aortic aneurysm (AAA) is a relatively common and life-threatening vascular disease characterized by the expansion of the infrarenal aorta. This expansion can lead to rupture, resulting in over 40,000 deaths annually in the United States ^1^. Despite this burden, there are no pharmacological therapies that are currently approved to treat AAA^2^.

Multiple genetic studies have identified risk loci associated with AAA susceptibility, with a recent genome-wide association study (GWAS) meta-analysis identifying over 100 independent loci ^3–7^. These studies, and others, have shown that AAA is a complex disease likely caused by a variety of interacting factors including inflammation and oxidative stress, along with extracellular matrix (ECM) and lipid metabolism dysfunction ^8^. Additionally, many of the loci associated with AAA encode proteins found in blood plasma ^3^.

Mendelian randomization (MR) is a statistical technique that utilizes genetic variation to infer potentially causal relationships between exposures and outcomes ^9^. In a manner analogous to randomization in a randomized control trial, MR leverages the random assortment of genetic variants at conception, as described by Mendel’s laws of inheritance, to help circumvent issues of confounding and reverse causation that can affect observational studies. By identifying genetic variants associated with an exposure of interest, MR can assess whether those variants are also associated with an outcome, allowing one to measure the potential causal effect of the exposure on the outcome. This approach has been successfully used to identify therapeutic targets for disease ^10,11^. Importantly, drugs supported by genetic evidence (such as MR) are 2-3 times more likely to be approved ^12^.

Recent advances have allowed for the quantification of thousands of plasma proteins and the identification of genetic variants associated with their levels (pQTLs) ^13,14^. Under certain assumptions, MR allows one to use this data to assess the causal effect of plasma protein levels on AAA. Given the proteome-wide nature of this data, MR can be utilized as a high-throughput computational screen to identify possible therapeutic targets.

In the framework of MR, we treated plasma protein levels as the exposures and AAA as the outcome. More specifically, we used pQTLs identified in two large-scale plasma-proteomics studies to generate instrumental variables for plasma protein levels ^13,14^. We then combined this with summary statistics from a recent GWAS meta-analysis for AAA ^3^ to run a plasma-proteome-wide MR screen to identify blood proteins with putative causal effects on the formation of AAA.

## Methods

### Initial MR analysis

Causal inference was performed using two-sample Mendelian randomization (MR). The three core assumptions underlying MR are: 1) the genetic variants used as instrumental variables (IVs) are associated with the exposure, 2) the IVs are only associated with the outcome through the exposure (and not through another factor), and 3) the IVs are not associated with any confounders that influence the exposure and the outcome ^10,15^. In practice, the first assumption can be met by using variants that are significantly associated with the exposure, as determined by a GWAS. The second and third assumptions are more difficult to satisfy. However, if the exposures are gene expression levels or protein levels, one can use *cis* genetic variants to minimize pleiotropy and maximize biological plausibility. Additionally, directional pleiotropy can be tested using MR-Egger regression ^16^. A significant intercept term could indicate directional pleiotropy and a violation of the second assumption.

For our analysis, plasma protein levels were taken to be the exposure, and AAA was taken to be the outcome. AAA summary statistics were obtained from (GWAS) meta-analysis including 39,221 individuals with and 1,086,107 individuals without AAA from 14 cohorts (AAAgen) ^3^. The study comprised 37,214 of European (EUR) ancestry and 2,007 of African (AFR) ancestry. For exposure data, we used publicly available protein quantitative trait loci (pQTLs) identified in two large-scale plasma-proteomics studies (deCODE and UKB-PPP), to generate genetic instruments ^13,14^. To our knowledge, none of the individuals used to generate the exposure data were part of the GWASs that comprised the AAA meta-analysis.

Variants were chosen as instrumental variables if their nominal p-values achieved genome-wide significance (p<5e-8). To reduce the risk of pleiotropic effects, we only considered *cis* variants located within 500 kb of their respective genes. We performed linkage disequilibrium (LD) clumping using the *ld_clump* function from the ‘R’ package ‘ieugawsr’ (version 1.0.0) ^17^. To allow more instruments per protein we relaxed the maximum LD R-squared threshold to 0.1. Otherwise, default argument values were used. We performed inverse-variance weighted MR using the *mr_ivw* function from the ‘R’ package ‘MendelianRandomization’ (version 0.10.0) ^18^. In cases where there is only a single genetic variant per exposure, the *mr_ivw* function returns a Wald ratio. Due to our relaxed LD threshold for variants, we performed MR with the ‘correl’ argument set to ‘TRUE’ to allow for correlated instruments. The Benjamini-Hochberg procedure was used to calculate a false discovery rate (FDR). Results were deemed significant at a threshold of FDR<0.05 in each dataset. For all initially significant proteins with at least 3 instrumental variables, we performed MR-Egger using the ‘MendelianRandomization’ function *mr_egger*. We followed STROBE-MR guidelines in reporting our findings^19^.

### Initial colocalization analysis

We performed Bayesian colocalization between plasma protein levels and AAA using the *coloc.abf* function from the ‘R’ package ‘coloc’ (version 5.2.3) ^20^. Default prior probabilities were used: p_1_=1e-4, p_2_=1e-4, p_12_=1e-5, where p_1_ and p_2_ are the prior probabilities that a variant is associated with trait 1 (plasma levels) and trait 2 (AAA) respectively. p_12_ is the prior probability that the variant is associated with both traits. We utilized all variants where summary statistics were available within 500 kb of genes encoding the proteins of interest. A posterior probability of a shared causal variant (H_4_) >0.7 was used as strong evidence of colocalization.

### Bi-directional MR

Bi-directional MR was performed to examine the possibility that genetic liability for AAA was causal for changes in plasma protein levels. We used the same approach as in our initial MR analysis, this time using genetic liability for AAA as the exposure and predicted plasma levels of our initially significant proteins as the outcome. However, all genome-wide significant (p<5e-8) variants for AAA were used as instruments (rather than only the significant *cis* variants used in the forward analysis).

### GO enrichment

GO analysis was performed using ‘Enrichr’^21^. Our gene set of interest consisted of the 25 genes/proteins identified as significant in the MR analysis (FDR<0.05) that were also supported by colocalization (posterior probability H_4_ >0.7). As a background, we used the set 2783 genes for which we were able to generate instruments and perform MR. Within our set of interest, we calculated the enrichment of ‘Reactome 2022’ pathways and ‘GO Cellular Component’ gene sets.

### Protein-Protein Interactions

Analysis was performed using ‘STRING’ ^22^. STRING is a database containing experimentally demonstrated and computationally predicted protein-protein interactions (PPIs). One can use this database to determine if the number of PPIs observed in a gene set of interest are more numerous than expected by chance. For our analysis, the gene set of interest and the background were identical to the GO enrichment analysis.

### Gene expression

Median TPM values across tissues were obtained from GTEx Portal (GTEx v8) ^23^. Expression levels were analyzed in vascular and endocrine tissues to account for plausible origins of plasma ECM proteins. For each of the 8 ECM proteins shown in **Figure 3C**, the median TPM levels within each tissue were Log_2_(TPM + 1) transformed. Those values were then divided by the Log_2_(TPM + 1) values in the tissue with the highest expression (out of the 18 tissues analyzed).

### Colocalization of eQTLs/sQTLs with pQTLs and AAA

Bayesian multi-trait colocalization was performed using the *hyprcoloc* function from the ‘R’ package ‘HyPrColoc’ (version 1.0) ^24,25^. Using summary statistics, this function allows one to calculate the posterior probability that all traits share a causal variant (similar to pp H_4_ in 2-trait colocalization). We used default parameters and prior values. Re-analyzed GTEx aortic tissue sQTL and eQTL summary statistics were obtained from ‘eQTL Catalogue’^26^. All available variants within 500 kb of our genes of interest were used for the analysis.

## Results

### MR identifies 90 plasma proteins associated with AAA

We used two-sample MR to estimate the effects of plasma protein levels on the genetic liability for AAA. We tested a total of 2,783 unique plasma proteins, with 1,701 from the deCODE dataset and 1,957 from the UKB dataset, for possible causal relationships with AAA using two-sample MR. The number of genetic variants used as instruments per protein ranged from 1 to 197, with a median of 10 variants for the deCODE data and 15 variants for the UKB data.

We found 52 plasma proteins significantly associated with AAA (FDR<0.05) using instruments from the deCODE dataset and another 50 proteins associated with AAA using instruments from the UKB dataset. (**Figure 1, Supplementary Table 1**). Overall, 90 unique proteins were found to be significantly associated with AAA. Of the 52 significant proteins found in the deCODE dataset, 23 were replicated in the UKB dataset (concordant direction of effect and nominal p-value<0.05). Conversely, 17 out of 50 proteins identified in the UKB dataset were replicated in the deCODE dataset. Notably, only 31/52 (60%) and 19/50 (38%) of the significant proteins found using instruments from the deCODE and UKB-PPP datasets respectively, had instruments in the other dataset. MR-Egger regression on plasma proteins significantly associated with AAA found no evidence of substantial directional pleiotropy (**Supplementary Table 2**).

**Figure 1.**
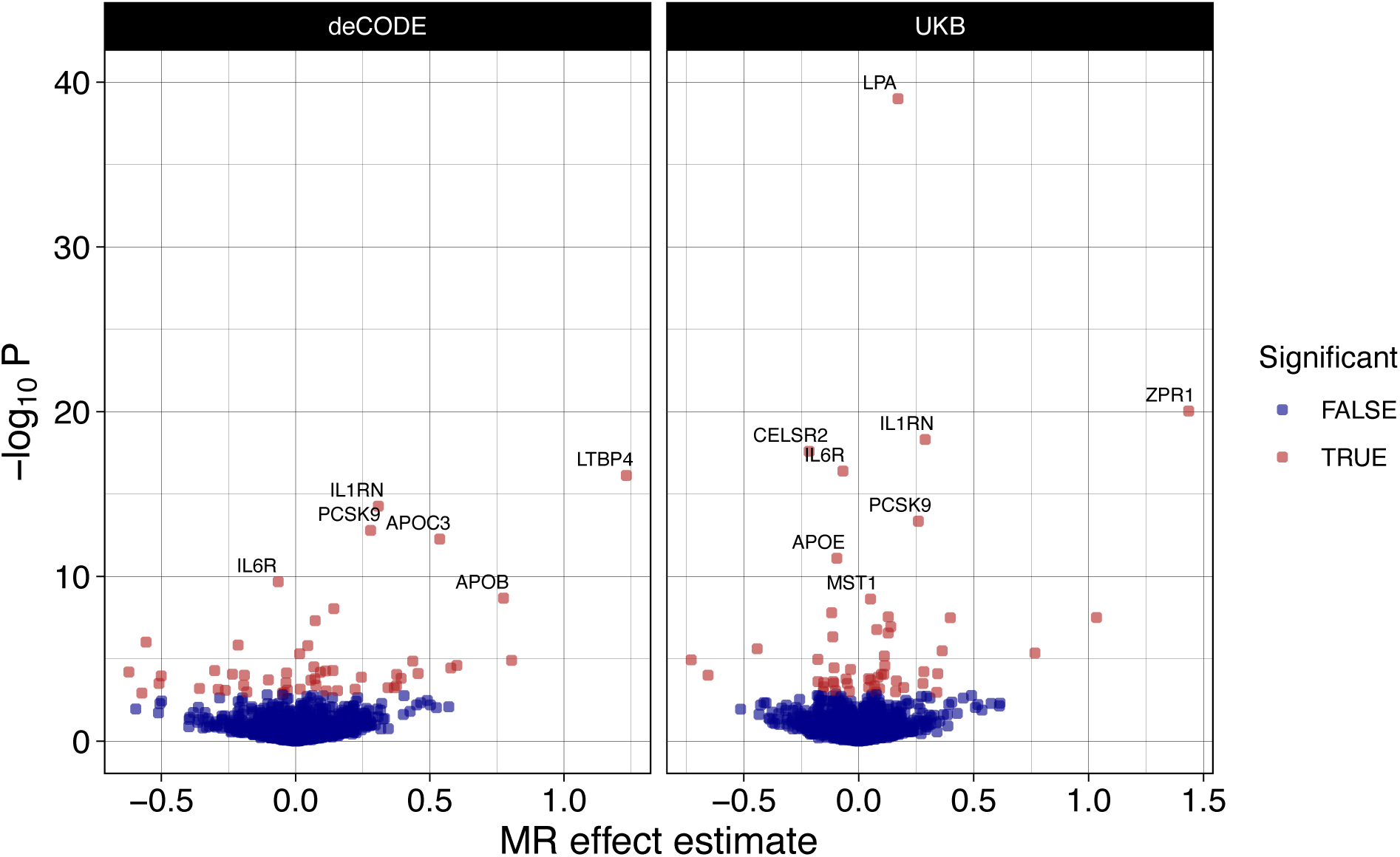
Mendelian randomization identifies 90 proteins associated with AAA. Instruments were generated and associations were tested for 2783 plasma proteins across the deCODE and UKB proteomics studies. The effect estimate is the natural logarithm of the odds ratio. Significance was determined at FDR<0.05 in each dataset. Proteins with p-adj<1e-6 are labeled.

### Colocalization provides additional support for a causal association between plasma protein levels and AAA

To provide additional support for the association between genetically determined levels of plasma proteins and genetic liability we tested for evidence of colocalization between pQTL and their cognate AAA risk loci ^27^. This approach can be helpful for distinguishing putative causal associations from those confounded by linkage disequilibrium ^28^. We used all *cis* variants within 500 kb of their respective genes to perform colocalization.

For 25 (27%) of the 90 proteins whose genetically determined plasma levels associated with genetic liability to AAA there was evidence of colocalization (posterior probability H4 >0.7), indicating that the plasma levels of these proteins and AAA likely share the same causal variants (**Figure 2, Supplementary Table 3**). Among those supported by both MR and colocalization were previously experimentally validated proteins such as PCSK9 (OR 1.3; 95%CI 1.2-1.4; P<1e-10), LTBP4 (OR 3.4; 95%CI 2.6-4.6; P<1e-10) and COL6A3 (OR 0.6; 95%CI 0.5-0.7; P<1e-6).

**Figure 2.**
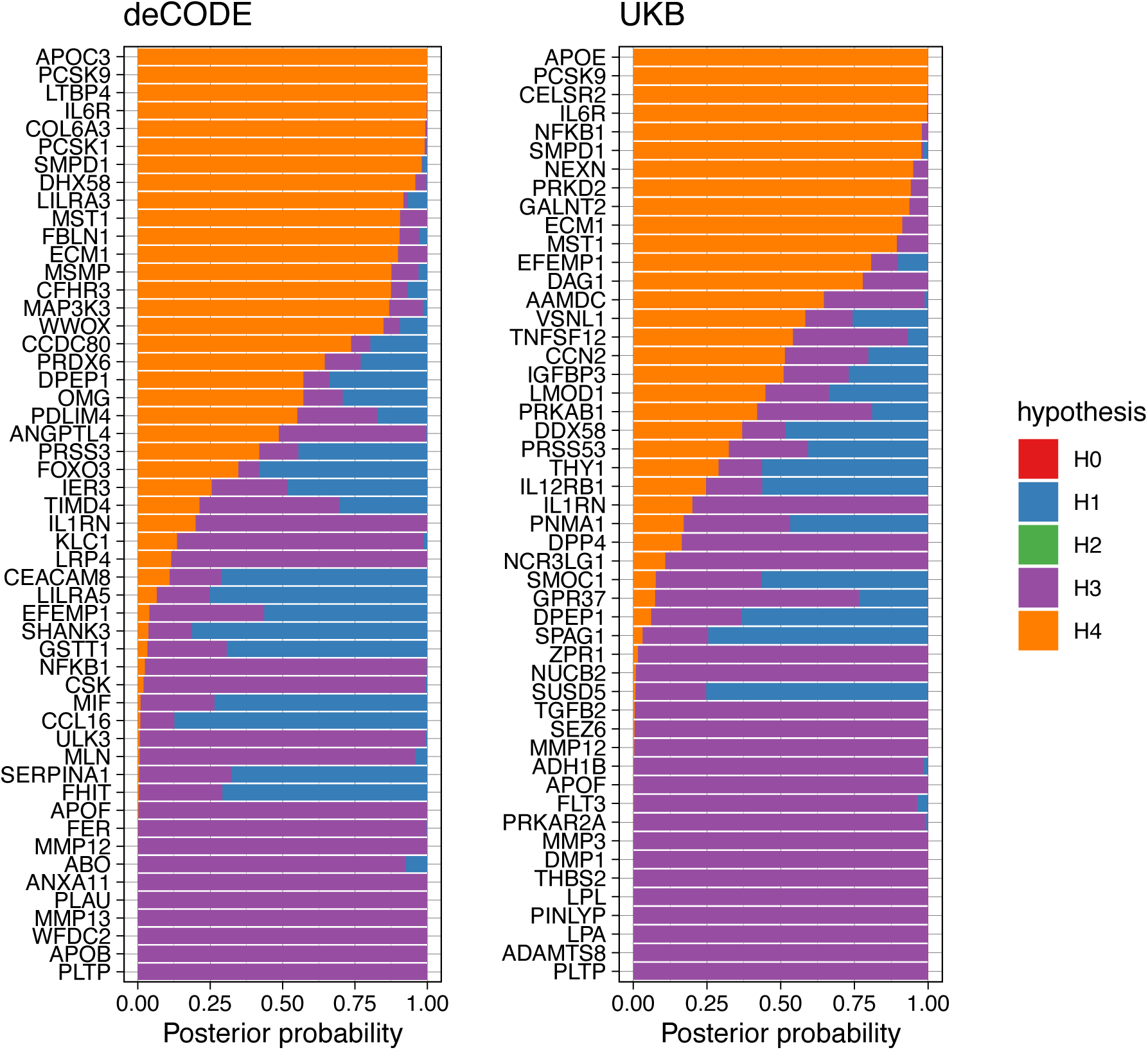
Colocalization analysis provides additional evidence for a causal association between protein levels and AAA. Bayesian colocalization results for all plasma proteins with significant associations with AAA (FDR<0.05). The hypotheses are as follows: H0) no causal variant for either trait, H1) causal variant for plasma levels only, H2) causal variant for AAA only, H3) two distinct causal variants, and H4) one shared causal variant for plasma levels and AAA. 25 distinct plasma proteins had posterior probability for H4>0.7, showing that the plasma protein levels and AAA likely share causal variants

### Putatively causal plasma proteins are enriched for proteins found in ECM

Among plasma proteins supported by both MR and colocalization, gene ontology (GO) enrichment analysis of Reactome pathways found an overrepresentation of proteins associated with elastic fibers (OR 29; p<1e-3), elastic fiber formation (OR 22; p<1e-3), and HDL remodeling (OR 80; p<1e-3) (**Figure 3A**) ^29^. GO analysis of cellular components revealed an enrichment of proteins found in collagen-containing extracellular matrix (ECM, OR 7.8; P<1e-4), chylomicrons (OR 48; P=1.6e-3), and endolysosomes (OR 34; P=2.7e-3) (**Figure 3A**). While our results were consistent with the current understanding of AAA as a disease influenced by ECM and lipid metabolism dysregulation ^8^, the fact that the ECM signal was generated from QTL derived from plasma measurement of proteins was not expected. To gain additional insights into the biology of these proteins, we investigated their protein-protein interactions (PPIs) (**Figure 3B**). Consistent with our GO analysis, we found that two clusters emerged: a cluster consisting of proteins involved in lipid metabolism and inflammation, and another cluster consisting of ECM proteins. The 24 genes included in this analysis (PPI data could not be found for LILRA3) had 16 known interactions among them, which was significantly higher than expected by chance (expected=9, p=0.015).

**Figure 3.**
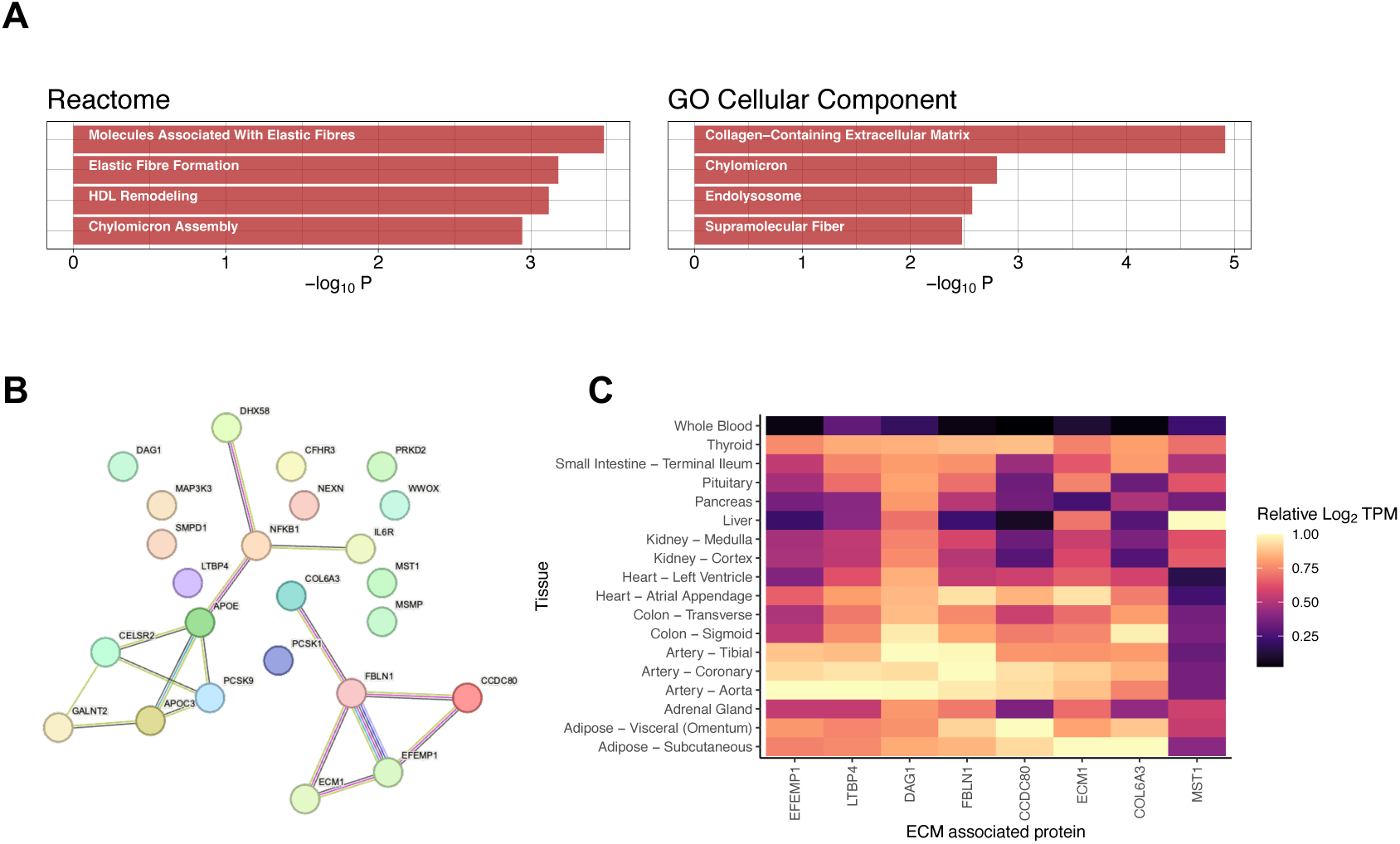
Biological properties of proteins associated with AAA. A) The 25 proteins supported by MR and colocalization were tested for significant overrepresentation of pathways and GO terms. The most significant Reactome pathway was “Molecules Associated with Elastic Fibers (OR 29; P<1e-3), while the most significant GO Cellular Component term was “Collagen-Containing Extracellular Matrix” (OR 7.8; P<1e-4). All terms and pathways shown were significant at FDR<0.05. B) Protein-protein interactions (PPIs) among proteins supported by both MR and colocalization. The PPIs were identified in the STRING database. An edge color indicates a particular type of evidence for each PPI (e.g. experimentally determined, co-expression, protein homology etc.). C) The expression levels of ECM associated proteins identified by MR and colocalization across tissues. Expression levels are shown relative to expression levels in the tissue with highest expression (among tissues shown). Median mRNA levels of the genes (TPM) in each tissue were Log_2_(TPM + 1) transformed and divided by the Log_2_(TPM + 1) transformed median TPM of the tissue with maximal expression. A value of 1 indicates that the tissue has the highest expression level for that gene out of the 18 tissues analyzed. Several of the ECM proteins show some of their highest expression levels in aortic tissue.

To help determine the origin of the AAA-associated ECM proteins we identified in blood plasma, we examined gene expression data from endocrine and blood-adjacent tissues found in the Genotype-Tissue Expression Project (GTEx) ^23^. We found that the proteins LTBP4, FBLN1, EFEMP1, and DAG1 had their highest mRNA levels in aortic or arterial tissues (**Figure 3C**). Additionally, CCDC80 and ECM1 had some of their highest expression levels in aortic tissue. These results indicate that the AAA-associated ECM proteins we identified in plasma plausibly originated in the aortic wall.

### Bi-directional MR shows that AAA is not likely to be causal for the changes in plasma levels of ECM proteins

To examine the possibility that AAA was causing the levels of ECM proteins to change in blood plasma we performed a second MR experiment, this time with genetic liability to AAA acting as the exposure and predicted plasma levels of each protein as the outcome (bi-directional MR). We performed bi-directional MR on all 90 proteins that we prioritized in our initial MR analysis. After accounting for multiple testing, only 6.7% (6/90) were putatively affected by genetic liability to AAA (FDR<0.05) (**Supplementary Table 4**). Notably, none of the ECM proteins appeared to have their circulating levels influenced by genetic susceptibility to AAA. As a complementary analysis, we used MR Steiger, which allows one to test whether an instrumental variable is more strongly associated with the exposure than the outcome – indicating that the exposure likely caused the outcome and not the reverse. We found that every instrumental variable used for proteins significantly associated with AAA had the appropriate directionality (**Supplementary Table 5**), further showing that AAA was unlikely to cause the observed changes in plasma protein levels.

### Variants affecting the expression and splicing of ECM proteins in the aortic wall may be causal for AAA

To gain insight into the biological mechanism by which genetic variation may lead to changes in ECM proteins in the aorta, and subsequently changes in plasma levels of these proteins, we examined expression quantitative trait loci (eQTLs) and splicing quantitative trait loci (sQTLs) in aortic tissue from GTEx. We performed a Bayesian colocalization analysis to determine whether the variants altering gene expression (mRNA levels or splicing) were also the same variants leading to changes in plasma levels and AAA. We considered all ECM proteins with expression TPM>10 in aortic tissue. We found that the eQTL for the ECM protein COL6A3 colocalized with its plasma pQTL and AAA with a posterior probability of 0.84 (**Figure 4**). At the putative causal variant, as determined by HyPrColoc ^24,25^, (GRCh38, chr2:237315312 A/G; rs11677932) the allele associated with increased aortic expression (G), was also associated with increased plasma levels and decreased risk of AAA.

**Figure 4.**
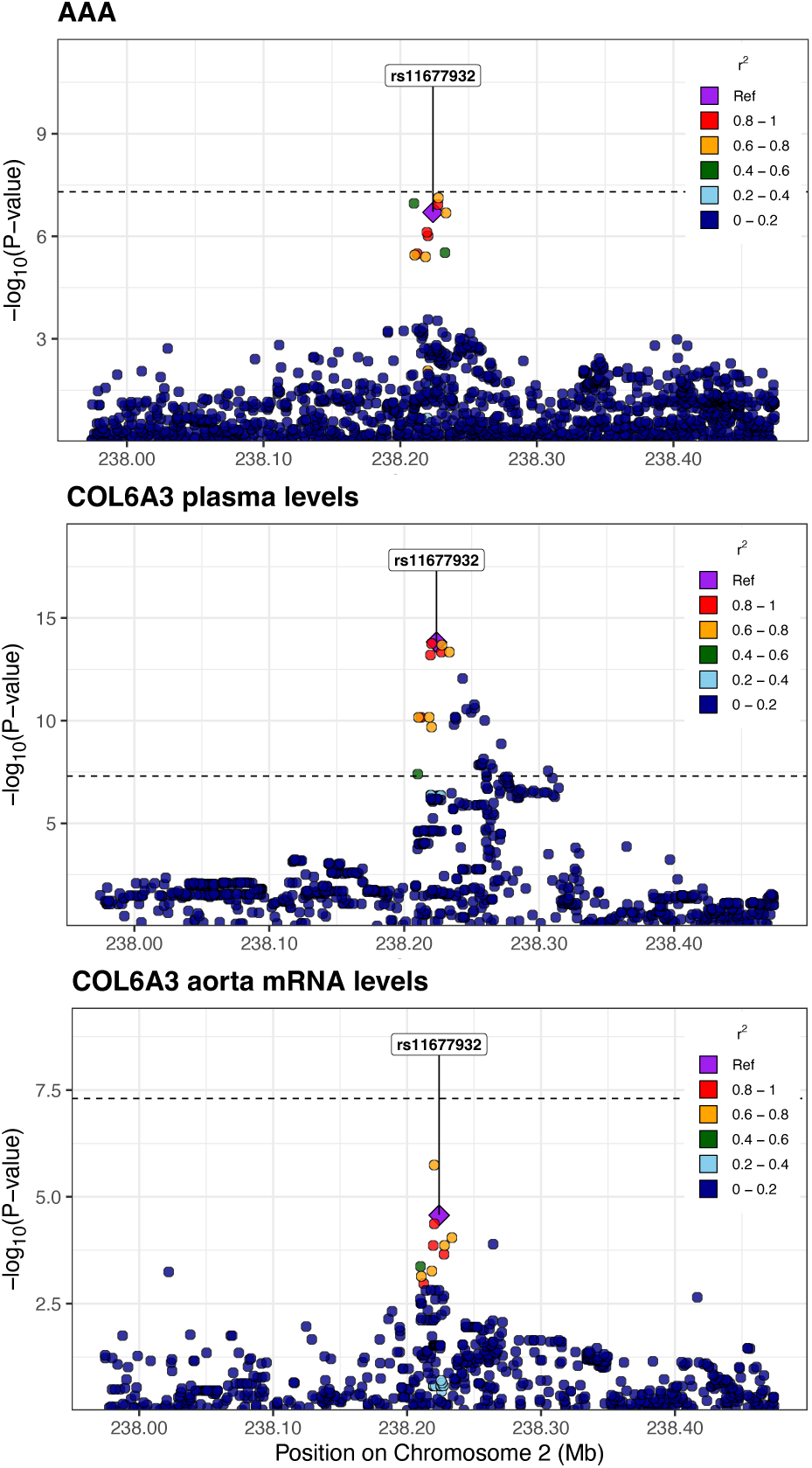
Regional association plots for COL6A3. Plots demonstrating the colocalization between variants associated with mRNA levels of COL6A3 in aorta, plasma levels of COL6A3, and AAA. The posterior probability for the colocalization of all 3 traits is 0.84.

The aptamer (used to measure plasma levels) for COL6A3 binds to the C-terminus, which is cleaved to a peptide called endotrophin ^30–32^. Given that COL6A3 had the highest expression levels in subcutaneous adipose tissue (in GTEx), and that plasma endotrophin has been linked to adipose tissue, we examined the possibility that the plasma COL6A3 originated in adipose tissue rather than aortic tissue. Hence, we performed an eQTL-pQTL-AAA colocalization analysis using eQTL data from subcutaneous adipose tissue. We found that the three signals did not colocalize (posterior probability = 6e-4), further adding evidence that increased plasma levels of COL6A3/endotrophin are caused by expression changes in aortic tissue.

We then examined sQTLs, and found that for LTBP4, at the putative causal variant (GRCh38, chr19: 40593595 A/T; rs112009052), the allele (T) associated with increased splicing out of an intron (GRCh38 chr19: 40611394:40611859) was associated with decreased transcription of a corresponding truncated transcript which does not use that splice junction pair (ENST00000243562). This allele was also associated with increased LTBP4 plasma levels and increased liability for AAA. The posterior probability for the colocalization of those four traits (sQTL, transcript-level-QTL, pQTL, and AAA) was 0.89 (**Figure 5**).

**Figure 5.**
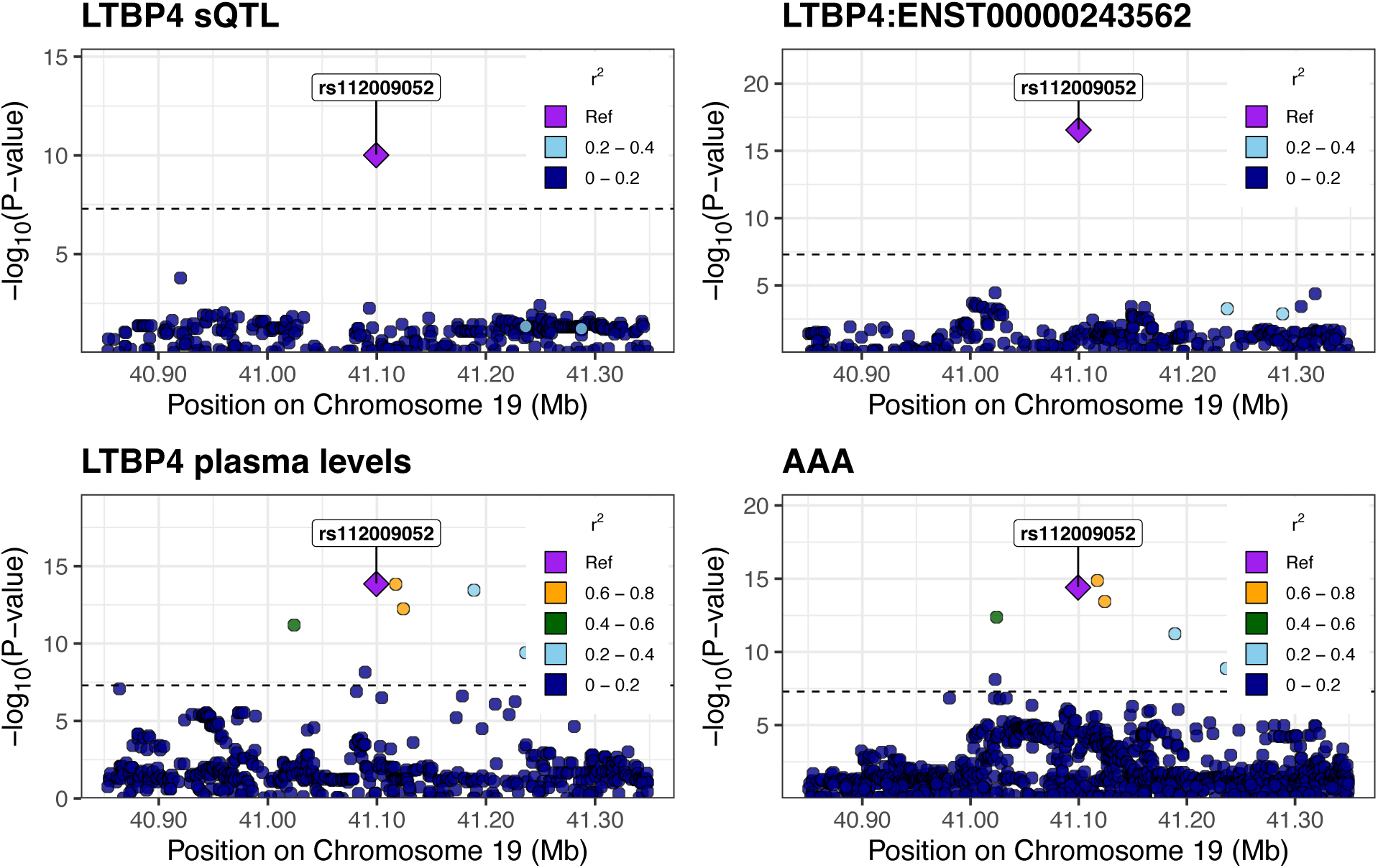
Regional association plots for LTBP4. Regional association plots demonstrating colocalization of sQTL and pQTL variants for LTBP4, with variants associated with AAA. The posterior probability for the colocalization of all 4 traits is 0.89.

To further investigate whether the expression or splicing changes cause changes in circulating protein levels, we performed MR. We used the putative causal variants identified by HyPrColoc as instruments, with eQTL (for COL6A3) or sQTL (for LTBP4) data as the exposure and pQTL data as the outcome. Our analysis revealed that COL6A3 aortic expression levels were significantly associated with COL6A3 plasma levels (OR 1.37, 95% CI 1.26-1.48, P=1.5e-14; Wald ratio). Similarly, LTBP4 splicing in the aorta was significantly associated with LTBP4 plasma levels (OR 1.15, 95% CI 1.11-1.20, P=1.4e-14; Wald ratio). These findings support the hypothesis that changes in expression or splicing in the aortic wall may drive alterations in plasma protein levels.

## Discussion

We performed proteome-wide MR to identify plasma proteins that are potentially causal in the development of AAA. Our analysis identified 90 distinct proteins associated with AAA. Of those, 25 were supported by colocalization analysis, indicating that the circulating protein levels and AAA shared causal variants. Unexpectedly, we found that the putatively causal proteins were enriched for proteins typically found in ECM, with several of the ECM proteins having their highest expression levels in aortic tissue.

Prior research has indicated that increased expression of matrix metalloproteinases (MMPs) directly leads to aneurysmal growth in AAA ^33^. MMPs are thought to induce aneurysmal growth through the breakdown of ECM proteins and thus the weakening of the aortic wall. Given this relationship, we investigated whether genetic liability for AAA might be causal for increased levels of circulating ECM proteins. However, our bi-directional MR and MR Steiger analyses demonstrated a lack of evidence for that hypothesis.

To examine the relationship between the aortic expression and the circulating levels of ECM proteins we performed a colocalization analysis. We sought to see if the variants responsible for changes in mRNA levels were also causal for changes in plasma levels and AAA. We examined all ECM proteins expressed in the aorta, and found that the eQTL colocalized (posterior probability = 0.84) with the pQTL and AAA variants for the protein COL6A3. All other eQTL-pQTL-AAA colocalization probabilities were below 0.5 for ECM proteins. We found additional support for the hypothesis that increased COL6A3 expression in the aorta causes an increase in COL6A3 plasma levels using MR.

The aptamer used to measure levels of COL6A3 in the deCODE proteomics dataset binds the C-terminal of COL6A3 ^13^. The C-terminal of COL6A3 gets cleaved into a peptide known as endotrophin, which can be found in the bloodstream ^30,31^. While expression data suggests that endotrophin may largely originate from subcutaneous adipose tissue, our colocalization analysis supports an alternative hypothesis: plasma endotrophin may primarily originate in aortic or vascular tissue.

Interestingly, our findings on endotrophin’s relationship with AAA contrast with its associations with other cardiovascular conditions. Clinical, functional, and computational studies have linked endotrophin to inflammation, insulin resistance, coronary artery disease (CAD) and heart failure (HF) ^30–32^. Notably, increased circulating levels of endotrophin were associated with increased risk of CAD and HF. However, our results suggest that increased plasma endotrophin may be protective against AAA. We hypothesize that this apparent protective effect against AAA might be related to endotrophin-induced insulin resistance, as insulin resistance has previously been associated with reduced rates of AAA ^34^. However, this relationship requires further investigation to establish causality and understand the underlying mechanisms.

To gain further insight into whether expression-level changes in the aortic wall were possibly responsible for AAA, we performed colocalization analysis between sQTLs, pQTLs, and AAA. Among all ECM proteins expression in aortic tissue we found that a sQTL for LTBP4 colocalized with its plasma pQTL and a significant locus for AAA. Furthermore, MR analysis provided additional evidence supporting the hypothesis that increased LTBP4 splicing in the aorta leads to an increase in LTBP4 plasma levels. These results suggest that splicing alterations in LTBP4 in the aortic wall may affect its plasma levels and AAA. Notably, across tissues, we found that LTBP4 is most highly expressed in aortic tissue (**Figure 3C**).

There are several different causal pathways by which ECM proteins, which are primarily expressed in the abdominal aortic wall, can lead to AAA while changing their circulating levels. The changes in the plasma levels themselves may or may not be causal in the formation of AAA. In the case that the plasma levels are not directly causal, and assuming that their origin is indeed the aorta, we can regard the plasma levels of these proteins as quantitative proxies for their ‘dysfunction’ within the aortic wall without invalidating the assumptions of MR. Prior research has demonstrated that LTBP4 is involved in the formation of elastic fibers ^35,36^ -- which is crucial to the integrity of the aorta ^33^. Therefore, it seems plausible that the original lesion in the causal pathway occurs in the aortic wall, and the elevated plasma levels of LTBP4 simply reflect its dysfunction in the aorta.

Given our findings, we propose a model for how ECM proteins found in plasma can cause AAA (**Figure 6**). Our findings suggest two plausible mechanisms linking genetic variation to abdominal aortic aneurysm (AAA). In one scenario, genetic factors alter the expression or splicing of extracellular matrix (ECM) proteins, leading to changes in the aortic wall. These alterations may directly contribute to AAA development while simultaneously causing ECM proteins to leak into the aortic lumen. Alternatively, the circulating ECM proteins themselves may be causative agents for AAA, independent of aortic wall changes. It’s also possible that a combination of both mechanisms is at play. Importantly, both hypotheses align with our findings and adhere to the assumptions of MR.

**Figure 6.**
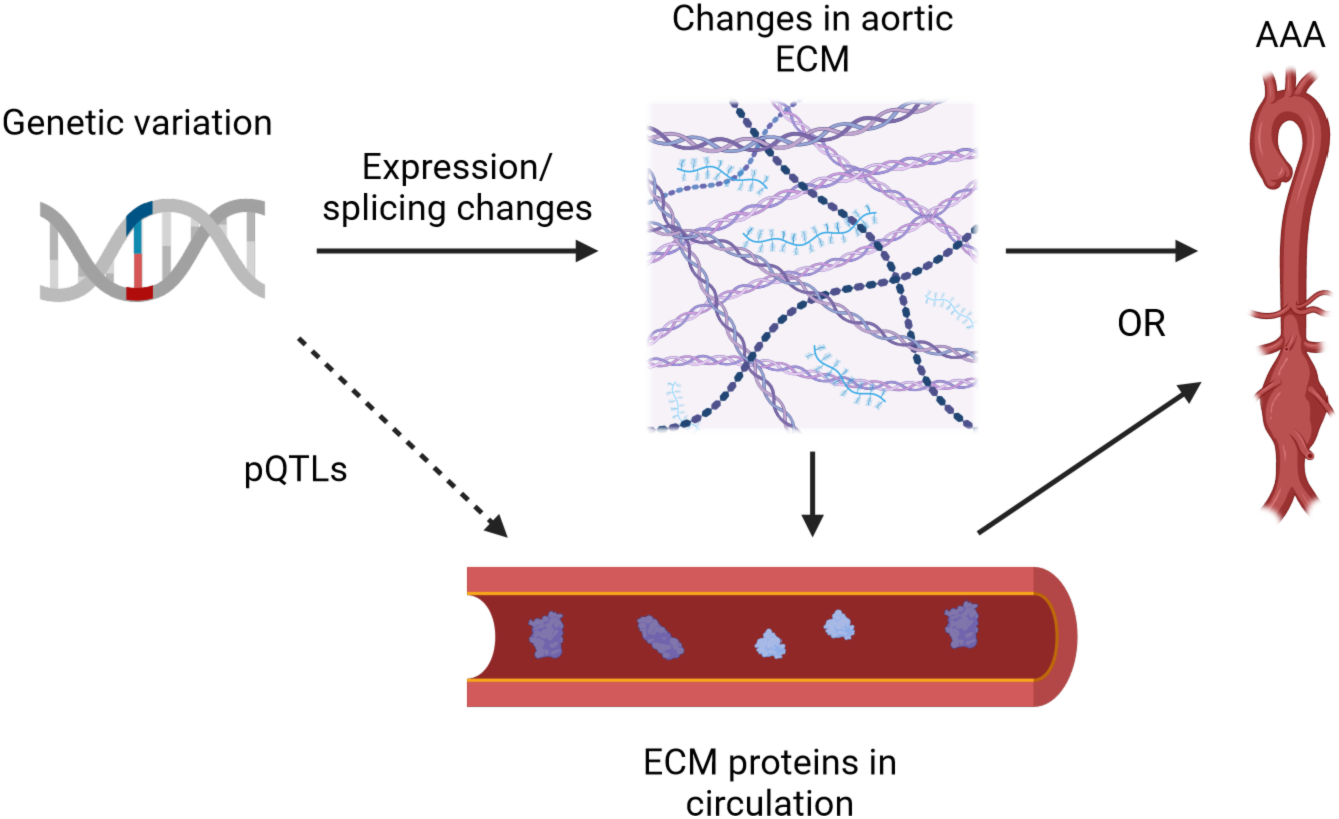
A causal model for AAA through ECM proteins. Genetic variation alters the expression or splicing of ECM proteins which leads to changes in the aortic wall. These aortic ECM changes lead to AAA, while simultaneously causing ECM proteins to leak out into the aortic lumen. Alternatively, the ECM proteins in circulation themselves cause AAA, rather than the aortic ECM changes. Both possibilities, or a combination of the two, are consistent with our findings and the assumptions of MR.

This study has several limitations despite our findings. Firstly, not all assumptions underlying Mendelian Randomization (MR) can be definitively proven. Although we attempted to minimize the risk of violation (see methods), two key assumptions remain unprovable: (1) that the instrumental variables (IVs) are associated with the outcome (AAA) only through the exposure (circulating protein levels), and (2) that the IVs are not associated with confounders affecting both the exposure and outcome. Additional experimental evidence is therefore necessary to prove causality.

A second limitation concerns our bi-directional MR analysis, which relied on both *trans*-pQTLs and *cis*-pQTLs to rule out the possibility that genetic liability for AAA causes changes in plasma ECM protein levels. However, the datasets used to discover these pQTLs are likely underpowered to detect trans-pQTLs, potentially affecting the robustness of this analysis.

Overall, our results highlight proteins and pathways with potential causal effects on AAA, providing a foundation for future functional experiments. These findings suggest a possible causal model for the formation of AAA through ECM proteins found in plasma.

## Data Availability

All data produced in the present study are available upon reasonable request to the authors.

## Acknowledgements

We want to thank the participants and investigators of the included GWASs and proteomics studies.

